## Supplementary material for "Determinants of COVID-19 vaccine uptake in the Netherlands: a nationwide registry-based study"

### S1 Long-term care recipients

To identify individuals receiving long-term care (LTC), two data sources were used. Firstly, start and end dates and types of LTC use were available within CBS. This is data from the CAK and includes LTC that is covered by the Long-term care act (WLZ) for which the LTC receiver pays a personal contribution to the CAK. An elaborate description of the database can be found (in Dutch) using the following URL: Gebwlztab: Personen Wlz-zorg (cbs.nl). Secondly, using the type of household variable in the personal records database (CBS) we were able to select individuals with 'institutional household' as type of household. A description of the personal records database and the variable type of household can be found (in Dutch) using the following URL: Gbapersoontab: Persoonskenmerken van personen in de BRP (cbs.nl)

In our analyses, we included three variables regarding LTC care distinguishing between residential and non-residential care and LTC for intellectual disabilities (residential and non-residential). For each variable, different selection methods were used:

1. LTC recipients, residential, nursing home: individuals were selected for this group if
    a. they received LTC care with profile VV5-8 (nursing and caring level 5-8) LTC and type of care 'care in kind'
    b. they lived in an institutional household according to the personal records database and received LTC with profile VV5-8
2. LTC recipients, residential, mentally impaired: individuals were selected for this group if
    a. they received LTC care with profile 'VG' (intellectually disabled) or 'LVG' (mildly intellectually disabled) and type of care 'care in kind'
    b. they lived in an institutional household according to the personal records database and received LTC with profile 'VG' or 'LVG'
3. LTC recipients, non-residential, mentally impaired: individuals were selected for this group if
    a. they received LTC care with profile 'VG' or 'LVG' and any type of care *other than* 'care in kind'
    b. they lived in any type of household *other than* institutional household according to the personal records database and received LTC with profile 'VG' or 'LVG'

### S2 Standard random forest and ROC analysis

The standard RF predictor was constructed and assessed using a dataset of 400,000 people. This provided estimates of prediction accuracy such as the **PMC** (probability of misclassification, i.e. the probability of predicting an individual's status incorrectly), the **sensitivity** (the probability of an individual's status being predicted as a '1', i.e. as vaccinated, when its status is indeed '1'), the **specificity** (the probability of an individual's status being predicted as a '0', i.e. as unvaccinated (or not having provided informed consent to share their status), when its status is indeed '0'), and the ranking of the predictor variables according to their importance, which was measured as the average increase in (worsening of) the PMC that results from the replacement of the value of a variable by a randomly chosen value. As expected, the standard RF yielded a very high sensitivity and a comparatively low specificity because there were far more vaccinated (80%) than unvaccinated people and the default prediction rule aims at minimizing the PMC (as opposed to the sensitivity or specificity).

Figure S2.1 shows the results of an ROC analysis based on the RF. The algorithm's ordinary prediction rule consists of predicting an individual's outcome as a 1 (vaccinated) if and only if the ratio of estimated probabilities of that individual being a 1 to it being a 0 conditionally on the predictor variables is ≥1. If we replace this inequality by $> c$ and we vary the value of the 'threshold' $c$ over positive values other than 1 we create a family of predictors, each with its own, adjusted prediction rule, and hence with its own performance characteristics. In particular, by varying the value of the threshold we get a varying pair of sensitivity and fpr (false positive rate = 1 minus specificity) estimates, which constitute the ROC curve, shown in the left panel of Fig. 2. By looking at the performance indicators obtained by varying the threshold, as shown on the right panel of Figure S3.1, we can pick the value which yields similar estimates of sensitivity and specificity; in this case this is a c of 3.55 , with corresponding values of 70% sensitivity and specificity and an increased PMC of 30%, together with the area under the ROC curve, 0.76.  The value of c was used to adjust the first RF, leading to the second, 'refined' RF, whose performance and variable importance were estimated by making predictions on the test data.

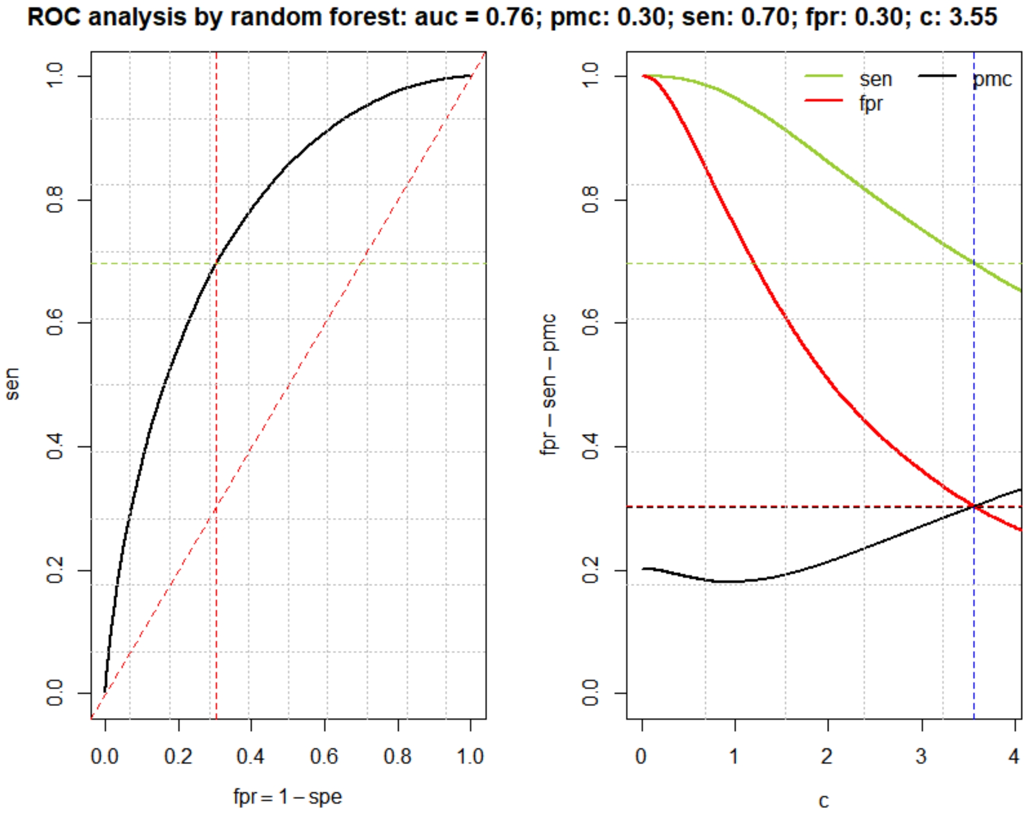

**Figure S2.1 ROC analysis by random forest.**

### S3 Vaccine uptake - Bivariate analyses

All reported tables exclude frequencies below ten and all numbers and percentages are rounded to the nearest ten, to avoid personally identifiable information.

#### Table S3.1 Vaccine uptake by age group

| | N | Uptake (N, %) |
|---|---|---|
| **Age group** | | |
| 18-35 | 4.012.330 | 2.683.820 (67) |
| 36-50 | 3.260.110 | 2.526.670 (78) |
| 51-66 | 3.845.610 | 3.339.460 (87) |
| 67-79 | 2.211.910 | 2.024.530 (92) |
| 80+ | 845.690 | 748.310 (88) |

#### Table S3.2 Vaccine uptake per determinant by age group

| | N | Uptake (N, %) |
|---|---|---|
| **Sex** | | |
| Male | 6.995.450 | 5.546.310 (79) |
| 18-35 | 2.037.610 | 1.345.890 (66) |
| 36-50 | 1.627.950 | 1.242.870 (76) |
| 51-66 | 1.922.600 | 1.674.810 (87) |
| 67-79 | 1.072.830 | 984.800 (92) |
| 80+ | 334.460 | 297.950 (89) |
| Female | 7.180.190 | 5.776.480 (80) |
| 18-35 | 1.974.720 | 1.337.930 (68) |
| 36-50 | 1.632.150 | 1.283.800 (79) |
| 51-66 | 1.923.010 | 1.664.650 (87) |
| 67-79 | 1.139.080 | 1.039.740 (91) |
| 80+ | 511.230 | 450.370 (88) |
| **Education level** | | |
| Primary education | 680.660 | 482.560 (71) |
| 18-35 | 115.660 | 59.860 (52) |
| 36-50 | 165.170 | 102.470 (62) |
| 51-66 | 243.680 | 185.030 (76) |
| 67-79 | 140.210 | 120.700 (86) |
| 80+ | 15.940 | 14.500 (91) |
| Prevocational secondary education-basic vocational programme (VMBO-b/k), lower secondary vocational training and assistant’s training (MBO-1) | 770.830 | 532.300 (69) |
| 18-35 | 256.470 | 120.450 (47) |
| 36-50 | 172.290 | 115.670 (67) |
| 51-66 | 235.650 | 198.120 (84) |
| 67-79 | 95.970 | 88.390 (92) |

| | | |
|---|---|---|
| 80+ | 10.460 | 9.670 (92) |
| Prevocational secondary education – theoretical and vocational programme (VMBO-g/t), the first three years of senior general secondary education (HAVO) and pre-university secondary education (VWO) | 385.580 | 273.660 (71) |
| 18-35 | 169.480 | 99.660 (59) |
| 36-50 | 73.070 | 49.550 (68) |
| 51-66 | 98.160 | 83.140 (85) |
| 67-79 | 41.490 | 38.150 (92) |
| 80+ | 3.370 | 3.150 (94) |
| Basic vocational training (MBO-2) and vocational training (MBO-3) | 1.428.730 | 1.005.180 (70) |
| 18-35 | 620.640 | 350.130 (56) |
| 36-50 | 331.250 | 237.140 (72) |
| 51-66 | 354.030 | 303.730 (86) |
| 67-79 | 113.790 | 105.750 (93) |
| 80+ | 9.020 | 8.450 (94) |
| Middle management and specialist education (MBO-4) | 1.470.350 | 1.094.650 (74) |
| 18-35 | 710.230 | 464.700 (65) |
| 36-50 | 379.970 | 293.240 (77) |
| 51-66 | 313.830 | 275.090 (88) |
| 67-79 | 63.620 | 59.100 (93) |
| 80+ | 2.690 | 2.530 (94) |
| Upper secondary education (HAVO/VWO) | 898.270 | 694.220 (77) |
| 18-35 | 568.380 | 421.100 (74) |
| 36-50 | 133.520 | 100.780 (75) |
| 51-66 | 156.840 | 136.040 (87) |
| 67-79 | 36.940 | 33.890 (92) |
| 80+ | 2.590 | 2.420 (94) |
| Hbo-, wo-bachelor | 2.055.220 | 1.700.130 (83) |
| 18-35 | 825.060 | 637.910 (77) |
| 36-50 | 674.610 | 561.610 (83) |
| 51-66 | 450.710 | 402.610 (89) |
| 67-79 | 98.960 | 92.460 (93) |
| 80+ | 5.880 | 5.550 (94) |
| Hbo-, wo-master, doctor | 1.204.530 | 1.067.060 (89) |
| 18-35 | 416.170 | 358.120 (86) |
| 36-50 | 438.950 | 387.290 (88) |
| 51-66 | 292.430 | 268.120 (92) |
| 67-79 | 53.690 | 50.400 (94) |
| 80+ | 3.290 | 3.140 (95) |
| Unknown | 5.281.490 | 4.473.030 (85) |
| 18-35 | 330.230 | 171.900 (52) |
| 36-50 | 891.270 | 678.930 (76) |
| 51-66 | 1.700.280 | 1.487.580 (87) |
| 67-79 | 1.567.240 | 1.435.700 (92) |
| 80+ | 792.460 | 698.910 (88) |

| **Country of origin** | | |
|---|---|---|
| The Netherlands | 10.660.430 | 9.045.520 (85) |
| 18-35 | 2.703.240 | 2.030.150 (75) |
| 36-50 | 2.269.290 | 1.874.450 (83) |
| 51-66 | 3.074.150 | 2.743.040 (89) |
| 67-79 | 1.883.420 | 1.746.340 (93) |
| 80+ | 730.340 | 651.540 (89) |
| Turkey | 333.090 | 177.880 (53) |
| 18-35 | 133.490 | 48.110 (36) |
| 36-50 | 110.090 | 67.880 (62) |
| 51-66 | 67.890 | 47.480 (70) |
| 67-79 | 16.800 | 11.160 (66) |
| 80+ | 4.830 | 3.250 (67) |
| Morocco | 294.480 | 119.200 (40) |
| 18-35 | 120.060 | 25.420 (21) |
| 36-50 | 94.790 | 43.960 (46) |
| 51-66 | 55.760 | 34.490 (62) |
| 67-79 | 17.900 | 11.770 (66) |
| 80+ | 5.980 | 3.560 (60) |
| Surinam | 304.370 | 191.400 (63) |
| 18-35 | 99.830 | 44.970 (45) |
| 36-50 | 83.550 | 52.020 (62) |
| 51-66 | 85.850 | 65.980 (77) |
| 67-79 | 28.710 | 23.400 (81) |
| 80+ | 6.420 | 5.020 (78) |
| The Dutch Caribbean | 136.230 | 70.510 (52) |
| 18-35 | 60.260 | 23.610 (39) |
| 36-50 | 35.650 | 18.850 (53) |
| 51-66 | 28.320 | 18.970 (67) |
| 67-79 | 10.310 | 7.820 (76) |
| 80+ | 1.690 | 1.270 (75) |
| Indonesia | 353.070 | 293.860 (83) |
| 18-35 | 42.620 | 29.610 (69) |
| 36-50 | 83.840 | 65.540 (78) |
| 51-66 | 131.790 | 114.410 (87) |
| 67-79 | 69.640 | 62.480 (90) |
| 80+ | 25.170 | 21.830 (87) |
| Other Africa | 227.900 | 134.870 (59) |
| 18-35 | 106.320 | 50.560 (48) |
| 36-50 | 66.990 | 43.690 (65) |
| 51-66 | 45.340 | 33.650 (74) |
| 67-79 | 7.960 | 6.010 (76) |
| 80+ | 1.290 | 970 (75) |
| Other Asia | 501.590 | 362.270 (72) |
| 18-35 | 230.590 | 149.920 (65) |
| 36-50 | 156.200 | 120.030 (77) |

| | | |
|---|---|---|
| 51-66 | 88.910 | 71.410 (80) |
| 67-79 | 21.830 | 17.830 (82) |
| 80+ | 4.070 | 3.080 (76) |
| Other America/Oceania | 195.140 | 142.310 (73) |
| 18-35 | 92.520 | 61.650 (67) |
| 36-50 | 55.130 | 41.810 (76) |
| 51-66 | 37.120 | 30.230 (81) |
| 67-79 | 8.980 | 7.490 (83) |
| 80+ | 1.400 | 1.140 (82) |
| Middle and eastern European countries within the EU | 304.230 | 137.150 (45) |
| 18-35 | 145.640 | 52.020 (36) |
| 36-50 | 105.500 | 50.800 (48) |
| 51-66 | 39.550 | 23.430 (59) |
| 67-79 | 11.200 | 9.020 (80) |
| 80+ | 2.340 | 1.890 (81) |
| GIPS countries | 142.440 | 96.850 (68) |
| 18-35 | 63.930 | 37.270 (58) |
| 36-50 | 42.550 | 30.870 (73) |
| 51-66 | 24.980 | 19.640 (79) |
| 67-79 | 7.530 | 6.280 (83) |
| 80+ | 3.450 | 2.800 (81) |
| Former or associated member states of the Commonwealth of Independent States | 46.170 | 29.150 (63) |
| 18-35 | 21.260 | 12.680 (60) |
| 36-50 | 14.430 | 9.240 (64) |
| 51-66 | 7.270 | 4.730 (65) |
| 67-79 | 2.710 | 2.170 (80) |
| 80+ | 510 | 340 (66) |
| Other countries of the EU | 516.720 | 408.360 (79) |
| 18-35 | 133.000 | 81.470 (61) |
| 36-50 | 97.580 | 75.350 (77) |
| 51-66 | 121.580 | 102.720 (84) |
| 67-79 | 109.180 | 99.540 (91) |
| 80+ | 55.390 | 49.280 (89) |
| Other European countries | 159.780 | 113.460 (71) |
| 18-35 | 59.570 | 36.400 (61) |
| 36-50 | 44.530 | 32.190 (72) |
| 51-66 | 37.120 | 29.290 (79) |
| 67-79 | 15.740 | 13.230 (84) |
| 80+ | 2.820 | 2.360 (83) |
| **Born in the Netherlands or abroad** | | |
| Born in The Netherlands with both parents born in The Netherlands | 10.660.430 | 9.045.520 (85) |
| 18-35 | 2.703.240 | 2.030.150 (75) |
| 36-50 | 2.269.290 | 1.874.450 (83) |

| | | |
|---|---|---|
| 51-66 | 3.074.150 | 2.743.040 (89) |
| 67-79 | 1.883.420 | 1.746.340 (93) |
| 80+ | 730.340 | 651.540 (89) |
| Born in the Netherlands with one parent born abroad | 735.910 | 570.470 (78) |
| 18-35 | 257.930 | 164.880 (64) |
| 36-50 | 158.800 | 121.540 (77) |
| 51-66 | 166.760 | 144.710 (87) |
| 67-79 | 107.170 | 98.760 (92) |
| 80+ | 45.260 | 40.590 (90) |
| Born in the Netherlands with two parents born abroad | 527.880 | 241.560 (46) |
| 18-35 | 337.560 | 113.810 (34) |
| 36-50 | 124.550 | 71.370 (57) |
| 51-66 | 48.760 | 41.090 (84) |
| 67-79 | 13.000 | 11.730 (90) |
| 80+ | 4.000 | 3.560 (89) |
| Born abroad with one parent born abroad | 84.660 | 66.410 (78) |
| 18-35 | 24.850 | 16.620 (67) |
| 36-50 | 19.300 | 14.510 (75) |
| 51-66 | 18.910 | 15.950 (84) |
| 67-79 | 15.080 | 13.610 (90) |
| 80+ | 6.520 | 5.720 (88) |
| Born abroad with two parents born abroad | 2.053.270 | 1.305.630 (64) |
| 18-35 | 653.310 | 331.700 (51) |
| 36-50 | 657.390 | 420.250 (64) |
| 51-66 | 508.020 | 369.170 (73) |
| 67-79 | 181.470 | 143.370 (79) |
| 80+ | 53.070 | 41.130 (78) |
| Born abroad with two parents born in the Netherlands | 113.490 | 93.220 (82) |
| 18-35 | 35.440 | 26.660 (75) |
| 36-50 | 30.780 | 24.560 (80) |
| 51-66 | 29.010 | 25.500 (88) |
| 67-79 | 11.760 | 10.720 (91) |
| 80+ | 6.500 | 5.780 (89) |
| **Socioeconomic position** | | |
| In employment | 6.984.360 | 5.618.950 (80) |
| 18-35 | 2.470.560 | 1.735.130 (70) |
| 36-50 | 2.324.820 | 1.911.280 (82) |
| 51-66 | 2.161.520 | 1.947.610 (90) |
| 67-79 | 26.030 | 23.650 (91) |
| 80+ | 1.430 | 1.280 (90) |
| Self-employed | 1.078.860 | 780.210 (72) |
| 18-35 | 253.550 | 135.610 (53) |
| 36-50 | 388.890 | 277.210 (71) |

| | | |
|---|---|---|
| 51-66 | 395.840 | 331.520 (84) |
| 67-79 | 38.140 | 33.820 (89) |
| 80+ | 2.440 | 2.070 (85) |
| Unemployment benefits (WW) | 103.080 | 72.400 (70) |
| 18-35 | 20.420 | 8.860 (43) |
| 36-50 | 33.820 | 21.960 (65) |
| 51-66 | 48.810 | 41.560 (85) |
| 67-79 | 20 | . |
| 80+ | . | . |
| Social assistance benefit | 416.890 | 233.240 (56) |
| 18-35 | 106.960 | 44.920 (42) |
| 36-50 | 141.830 | 76.580 (54) |
| 51-66 | 165.130 | 109.580 (66) |
| 67-79 | 1.960 | 1.480 (76) |
| 80+ | 1.020 | 680 (67) |
| Other benefits | 266.950 | 178.090 (67) |
| 18-35 | 117.520 | 68.330 (58) |
| 36-50 | 70.180 | 46.990 (67) |
| 51-66 | 78.100 | 61.820 (79) |
| 67-79 | 380 | 320 (83) |
| 80+ | 770 | 630 (82) |
| Disability benefit | 521.330 | 387.210 (74) |
| 18-35 | 55.310 | 25.850 (47) |
| 36-50 | 140.340 | 94.440 (67) |
| 51-66 | 325.650 | 266.890 (82) |
| 67-79 | 30 | . |
| 80+ | . | . |
| Pensioner | 3.429.200 | 3.106.120 (91) |
| 18-35 | 3.730 | 2.180 (59) |
| 36-50 | 17.420 | 12.620 (72) |
| 51-66 | 426.260 | 384.420 (90) |
| 67-79 | 2.142.670 | 1.963.720 (92) |
| 80+ | 839.120 | 743.180 (89) |
| Student | 832.380 | 612.810 (74) |
| 18-35 | 825.380 | 607.400 (74) |
| 36-50 | 5.730 | 4.360 (76) |
| 51-66 | 1.260 | 1.040 (82) |
| 67-79 | 20 | . |
| 80+ | . | . |
| Other/unknown | 542.590 | 333.770 (62) |
| 18-35 | 158.910 | 55.550 (35) |
| 36-50 | 137.080 | 81.240 (59) |
| 51-66 | 243.030 | 195.030 (80) |
| 67-79 | 2.660 | 1.480 (56) |
| 80+ | 910 | 470 (52) |

| | | |
|---|---|---|
| **Personal income (percentiles)** | | |
| 0 till 10 | 706.770 | 488.830 (69) |
| 18-35 | 447.550 | 312.420 (70) |
| 36-50 | 86.120 | 54.440 (63) |
| 51-66 | 139.530 | 112.140 (80) |
| 67-79 | 16.980 | 5.330 (31) |
| 80+ | 16.590 | 4.510 (27) |
| 10 till 25 | 2.033.890 | 1.582.610 (78) |
| 18-35 | 644.150 | 414.500 (64) |
| 36-50 | 240.810 | 161.310 (67) |
| 51-66 | 366.730 | 300.720 (82) |
| 67-79 | 603.840 | 548.200 (91) |
| 80+ | 178.360 | 157.880 (89) |
| 25 till 50 | 3.481.510 | 2.695.920 (77) |
| 18-35 | 864.710 | 519.310 (60) |
| 36-50 | 640.570 | 452.280 (71) |
| 51-66 | 811.500 | 668.440 (82) |
| 67-79 | 803.200 | 729.430 (91) |
| 80+ | 361.540 | 326.450 (90) |
| 50 till 75 | 3.485.790 | 2.836.700 (81) |
| 18-35 | 1.073.340 | 747.850 (70) |
| 36-50 | 832.310 | 654.310 (79) |
| 51-66 | 933.650 | 825.990 (88) |
| 67-79 | 501.880 | 473.180 (94) |
| 80+ | 144.620 | 135.370 (94) |
| 75 till 90 | 2.091.710 | 1.787.130 (85) |
| 18-35 | 515.240 | 398.710 (77) |
| 36-50 | 675.350 | 560.580 (83) |
| 51-66 | 696.980 | 633.000 (91) |
| 67-79 | 167.180 | 159.740 (96) |
| 80+ | 36.960 | 35.100 (95) |
| 90 till 100 | 1.533.870 | 1.365.600 (89) |
| 18-35 | 201.730 | 158.650 (79) |
| 36-50 | 616.310 | 538.930 (87) |
| 51-66 | 618.540 | 574.880 (93) |
| 67-79 | 82.640 | 79.150 (96) |
| 80+ | 14.640 | 13.990 (96) |
| Unknown | 842.090 | 566.020 (67) |
| 18-35 | 265.620 | 132.380 (50) |
| 36-50 | 168.650 | 104.820 (62) |
| 51-66 | 278.670 | 224.290 (80) |
| 67-79 | 36.190 | 29.510 (82) |
| 80+ | 92.980 | 75.010 (81) |
| **Household type** | | |
| One-person household | 3.075.860 | 2.344.020 (76) |

| | | |
|---|---|---|
| 18-35 | 878.160 | 569.020 (65) |
| 36-50 | 499.110 | 336.070 (67) |
| 51-66 | 728.590 | 584.370 (80) |
| 67-79 | 586.680 | 516.430 (88) |
| 80+ | 383.320 | 338.140 (88) |
| Unmarried couple without children | 1.257.950 | 988.890 (79) |
| 18-35 | 665.210 | 493.520 (74) |
| 36-50 | 209.100 | 157.400 (75) |
| 51-66 | 272.620 | 236.400 (87) |
| 67-79 | 94.100 | 86.260 (92) |
| 80+ | 16.910 | 15.310 (91) |
| Married couple without children | 3.372.970 | 3.059.750 (91) |
| 18-35 | 188.300 | 128.090 (68) |
| 36-50 | 190.400 | 153.840 (81) |
| 51-66 | 1.330.510 | 1.218.040 (92) |
| 67-79 | 1.350.010 | 1.270.760 (94) |
| 80+ | 313.760 | 289.020 (92) |
| Unmarried couple with children | 1.052.990 | 799.390 (76) |
| 18-35 | 374.200 | 242.570 (65) |
| 36-50 | 495.060 | 399.570 (81) |
| 51-66 | 171.380 | 147.210 (86) |
| 67-79 | 10.930 | 8.980 (82) |
| 80+ | 1.430 | 1.060 (74) |
| Married couple with children | 4.101.880 | 3.248.490 (79) |
| 18-35 | 1.392.250 | 966.940 (69) |
| 36-50 | 1.523.520 | 1.245.210 (82) |
| 51-66 | 1.071.830 | 940.260 (88) |
| 67-79 | 97.260 | 82.590 (85) |
| 80+ | 17.020 | 13.490 (79) |
| One-parent family | 965.810 | 628.550 (65) |
| 18-35 | 400.760 | 214.350 (53) |
| 36-50 | 295.490 | 202.700 (69) |
| 51-66 | 219.070 | 171.700 (78) |
| 67-79 | 32.920 | 26.080 (79) |
| 80+ | 17.580 | 13.720 (78) |
| Other household | 101.440 | 63.750 (63) |
| 18-35 | 65.660 | 38.610 (59) |
| 36-50 | 13.470 | 7.330 (54) |
| 51-66 | 14.380 | 11.160 (78) |
| 67-79 | 6.080 | 5.130 (84) |
| 80+ | 1.840 | 1.520 (82) |
| Institutional household | 246.740 | 189.960 (77) |
| 18-35 | 47.780 | 30.710 (64) |
| 36-50 | 33.960 | 24.570 (72) |
| 51-66 | 37.220 | 30.330 (81) |
| 67-79 | 33.930 | 28.290 (83) |

| | | |
|---|---|---|
| 80+ | 93.840 | 76.060 (81) |
| **Household car ownership** | | |
| Yes | 11.113.920 | 9.101.130 (82) |
| 18-35 | 2.848.490 | 1.929.370 (68) |
| 36-50 | 2.669.600 | 2.122.330 (79) |
| 51-66 | 3.283.050 | 2.904.110 (88) |
| 67-79 | 1.838.870 | 1.712.610 (93) |
| 80+ | 473.900 | 432.710 (91) |
| No | 3.061.720 | 2.221.670 (73) |
| 18-35 | 1.163.830 | 754.460 (65) |
| 36-50 | 590.500 | 404.340 (68) |
| 51-66 | 562.560 | 435.350 (77) |
| 67-79 | 373.040 | 311.920 (84) |
| 80+ | 371.790 | 315.600 (85) |
| **Employment sector** | | |
| Activities of household as employer, undifferentiated goods and service producing activities of household for own use | 31.030 | 21.420 (69) |
| 18-35 | 6.320 | 2.730 (43) |
| 36-50 | 9.150 | 6.050 (66) |
| 51-66 | 11.360 | 8.960 (79) |
| 67-79 | 3.790 | 3.320 (88) |
| 80+ | 410 | 360 (90) |
| Agriculture, forestry and fishery | 73.830 | 57.290 (78) |
| 18-35 | 29.310 | 20.080 (69) |
| 36-50 | 21.140 | 16.510 (78) |
| 51-66 | 21.310 | 18.780 (88) |
| 67-79 | 1.950 | 1.800 (93) |
| 80+ | 130 | . |
| Mining and quarrying | 6.910 | 5.820 (84) |
| 18-35 | 1.800 | 1.380 (77) |
| 36-50 | 2.910 | 2.450 (84) |
| 51-66 | 2.170 | 1.960 (90) |
| 67-79 | 40 | . |
| 80+ | . | . |
| Industry | 717.940 | 586.020 (82) |
| 18-35 | 205.920 | 145.140 (70) |
| 36-50 | 240.560 | 196.370 (82) |
| 51-66 | 265.370 | 238.770 (90) |
| 67-79 | 5.850 | 5.520 (94) |
| 80+ | 240 | 230 (94) |
| Electricity supply | 29.730 | 25.260 (85) |
| 18-35 | 8.870 | 6.780 (76) |
| 36-50 | 10.960 | 9.300 (85) |
| 51-66 | 9.610 | 8.910 (93) |
| 67-79 | 280 | 260 (94) |

| | | |
|---|---|---|
| 80+ | 10 | . |
| Water supply, sewage and waste management | 35.770 | 28.660 (80) |
| 18-35 | 9.270 | 6.130 (66) |
| 36-50 | 12.390 | 9.890 (80) |
| 51-66 | 13.900 | 12.440 (90) |
| 67-79 | 200 | 190 (94) |
| 80+ | . | . |
| Construction | 323.500 | 255.540 (79) |
| 18-35 | 109.010 | 72.450 (66) |
| 36-50 | 113.030 | 91.370 (81) |
| 51-66 | 98.230 | 88.700 (90) |
| 67-79 | 3.110 | 2.900 (93) |
| 80+ | 130 | . |
| Wholesale and retail trade | 1.123.120 | 881.150 (78) |
| 18-35 | 512.910 | 358.040 (70) |
| 36-50 | 326.750 | 267.110 (82) |
| 51-66 | 267.510 | 240.990 (90) |
| 67-79 | 15.310 | 14.410 (94) |
| 80+ | 640 | 600 (93) |
| Transportation and storage | 363.240 | 280.800 (77) |
| 18-35 | 113.780 | 71.740 (63) |
| 36-50 | 111.120 | 86.260 (78) |
| 51-66 | 128.980 | 113.990 (88) |
| 67-79 | 9.180 | 8.640 (94) |
| 80+ | 180 | 170 (94) |
| Accommodation and food services activities | 261.770 | 198.610 (76) |
| 18-35 | 163.730 | 116.810 (71) |
| 36-50 | 54.940 | 43.730 (80) |
| 51-66 | 40.580 | 35.700 (88) |
| 67-79 | 2.420 | 2.280 (94) |
| 80+ | 110 | . |
| Information and communication | 291.390 | 245.100 (84) |
| 18-35 | 138.630 | 110.630 (80) |
| 36-50 | 97.840 | 84.000 (86) |
| 51-66 | 53.790 | 49.420 (92) |
| 67-79 | 1.090 | 1.020 (93) |
| 80+ | 40 | . |
| Financial services | 264.170 | 230.000 (87) |
| 18-35 | 70.100 | 55.320 (79) |
| 36-50 | 103.530 | 90.310 (87) |
| 51-66 | 82.900 | 77.120 (93) |
| 67-79 | 7.010 | 6.650 (95) |
| 80+ | 620 | 590 (95) |
| Real estate activities | 65.700 | 55.230 (84) |
| 18-35 | 19.260 | 14.060 (73) |
| 36-50 | 23.850 | 20.370 (85) |

| | | |
|---|---|---|
| 51-66 | 20.720 | 19.020 (92) |
| 67-79 | 1.690 | 1.600 (94) |
| 80+ | 190 | 180 (93) |
| Professional scientific and technical activities | 515.870 | 440.230 (85) |
| 18-35 | 223.000 | 179.200 (80) |
| 36-50 | 166.110 | 143.960 (87) |
| 51-66 | 119.250 | 109.970 (92) |
| 67-79 | 7.230 | 6.850 (95) |
| 80+ | 280 | 260 (93) |
| Administrative and support service activities | 740.260 | 513.200 (69) |
| 18-35 | 367.700 | 223.950 (61) |
| 36-50 | 196.570 | 139.810 (71) |
| 51-66 | 163.090 | 137.330 (84) |
| 67-79 | 12.710 | 11.930 (94) |
| 80+ | 190 | 180 (94) |
| Public administration and defence | 529.030 | 455.280 (86) |
| 18-35 | 142.170 | 110.050 (77) |
| 36-50 | 183.370 | 157.170 (86) |
| 51-66 | 202.020 | 186.640 (92) |
| 67-79 | 1.450 | 1.400 (97) |
| 80+ | 20 | . |
| Education | 529.830 | 459.890 (87) |
| 18-35 | 185.110 | 149.120 (81) |
| 36-50 | 178.860 | 156.640 (88) |
| 51-66 | 162.420 | 150.880 (93) |
| 67-79 | 3.380 | 3.200 (95) |
| 80+ | 60 | . |
| Human health and social work activities | 1.348.300 | 1.113.670 (83) |
| 18-35 | 509.860 | 377.020 (74) |
| 36-50 | 402.870 | 340.760 (85) |
| 51-66 | 429.630 | 390.340 (91) |
| 67-79 | 5.870 | 5.480 (93) |
| 80+ | 80 | . |
| Arts, entertainment and recreation | 106.330 | 87.350 (82) |
| 18-35 | 49.420 | 37.710 (76) |
| 36-50 | 27.650 | 22.930 (83) |
| 51-66 | 27.540 | 25.070 (91) |
| 67-79 | 1.640 | 1.570 (96) |
| 80+ | 90 | 80 (89) |
| Other service activities | 116.860 | 94.510 (81) |
| 18-35 | 45.700 | 32.260 (71) |
| 36-50 | 35.600 | 29.780 (84) |
| 51-66 | 33.460 | 30.520 (91) |
| 67-79 | 2.000 | 1.880 (94) |
| 80+ | 90 | . |
| Activities of extraterritorial organisations and bodies | 1.090 | 800 (73) |

| | | |
|---|---|---|
| 18-35 | 220 | 150 (68) |
| 36-50 | 400 | 280 (71) |
| 51-66 | 460 | 360 (77) |
| 67-79 | 10 | . |
| 80+ | . | . |
| Other/unemployed/unknown | 6.699.980 | 5.286.970 (79) |
| 18-35 | 1.100.270 | 593.090 (54) |
| 36-50 | 940.520 | 611.630 (65) |
| 51-66 | 1.691.310 | 1.393.600 (82) |
| 67-79 | 2.125.700 | 1.943.610 (91) |
| 80+ | 842.190 | 745.040 (88) |
| **Urbanisation** | | |
| Not urbanised | 2.267.080 | 1.869.340 (82) |
| 18-35 | 534.040 | 373.250 (70) |
| 36-50 | 494.820 | 392.120 (79) |
| 51-66 | 721.400 | 632.820 (88) |
| 67-79 | 390.800 | 359.620 (92) |
| 80+ | 126.020 | 111.530 (88) |
| Hardly urbanised | 2.243.130 | 1.882.930 (84) |
| 18-35 | 538.900 | 384.790 (71) |
| 36-50 | 522.880 | 428.280 (82) |
| 51-66 | 648.070 | 578.700 (89) |
| 67-79 | 386.890 | 359.970 (93) |
| 80+ | 146.400 | 131.180 (90) |
| Moderately urbanised | 2.553.650 | 2.108.270 (83) |
| 18-35 | 635.300 | 436.490 (69) |
| 36-50 | 599.820 | 482.760 (80) |
| 51-66 | 726.450 | 644.130 (89) |
| 67-79 | 430.150 | 399.600 (93) |
| 80+ | 161.940 | 145.290 (90) |
| Strongly urbanised | 3.600.060 | 2.854.700 (79) |
| 18-35 | 969.800 | 621.040 (64) |
| 36-50 | 840.940 | 647.600 (77) |
| 51-66 | 970.810 | 842.780 (87) |
| 67-79 | 580.300 | 531.760 (92) |
| 80+ | 238.210 | 211.530 (89) |
| Extremely urbanised | 3.510.730 | 2.607.110 (74) |
| 18-35 | 1.333.780 | 868.060 (65) |
| 36-50 | 801.410 | 575.810 (72) |
| 51-66 | 778.710 | 640.920 (82) |
| 67-79 | 423.720 | 373.530 (88) |
| 80+ | 173.120 | 148.790 (86) |
| Unknown | 990 | 440 (44) |
| 18-35 | 500 | 180 (36) |
| 36-50 | 240 | 100 (41) |

| | | |
|---|---|---|
| 51-66 | 180 | 110 (61) |
| 67-79 | 60 | 40 (77) |
| 80+ | . | . |
| **Medical risk groups** | | |
| Low medical risk | 10.598.450 | 8.259.750 (78) |
| 18-35 | 3.577.840 | 2.385.010 (67) |
| 36-50 | 2.708.720 | 2.084.480 (77) |
| 51-66 | 2.743.830 | 2.367.020 (86) |
| 67-79 | 1.216.950 | 1.110.960 (91) |
| 80+ | 351.120 | 312.290 (89) |
| Intermediate medical risk | 3.216.720 | 2.749.800 (85) |
| 18-35 | 399.380 | 273.340 (68) |
| 36-50 | 488.490 | 389.270 (80) |
| 51-66 | 976.410 | 860.680 (88) |
| 67-79 | 891.360 | 819.930 (92) |
| 80+ | 461.090 | 406.570 (88) |
| High medical risk | 360.480 | 313.250 (87) |
| 18-35 | 35.110 | 25.480 (73) |
| 36-50 | 62.910 | 52.920 (84) |
| 51-66 | 125.370 | 111.760 (89) |
| 67-79 | 103.600 | 93.640 (90) |
| 80+ | 33.490 | 29.460 (88) |
| **Long term care recipients, residential, nursing home** | | |
| No | 14.066.530 | 11.232.240 (80) |
| 18-35 | 4.012.270 | 2.683.780 (67) |
| 36-50 | 3.259.660 | 2.526.300 (78) |
| 51-66 | 3.840.720 | 3.335.270 (87) |
| 67-79 | 2.188.690 | 2.004.750 (92) |
| 80+ | 765.200 | 682.160 (89) |
| Yes | 109.110 | 90.550 (83) |
| 18-35 | 60 | 50 (81) |
| 36-50 | 450 | 370 (82) |
| 51-66 | 4.890 | 4.190 (86) |
| 67-79 | 23.220 | 19.790 (85) |
| 80+ | 80.490 | 66.160 (82) |
| **Long term care recipients, residential, mentally impaired** | | |
| No | 14.100.410 | 11.260.710 (80) |
| 18-35 | 3.984.460 | 2.662.280 (67) |
| 36-50 | 3.242.510 | 2.511.740 (77) |
| 51-66 | 3.825.040 | 3.321.600 (87) |
| 67-79 | 2.204.090 | 2.017.940 (92) |
| 80+ | 844.310 | 747.140 (88) |
| Yes | 75.230 | 62.090 (83) |

| | | |
|---|---|---|
| 18-35 | 27.870 | 21.540 (77) |
| 36-50 | 17.600 | 14.930 (85) |
| 51-66 | 20.560 | 17.860 (87) |
| 67-79 | 7.820 | 6.590 (84) |
| 80+ | 1.390 | 1.170 (84) |
| **Long term care recipients, non-residential, mentally impaired** | | |
| No | 14.149.170 | 11.303.980 (80) |
| 18-35 | 3.993.510 | 2.670.900 (67) |
| 36-50 | 3.255.850 | 2.523.510 (78) |
| 51-66 | 3.843.000 | 3.337.370 (87) |
| 67-79 | 2.211.190 | 2.023.940 (92) |
| 80+ | 845.620 | 748.260 (88) |
| Yes | 26.470 | 18.820 (71) |
| 18-35 | 18.820 | 12.920 (69) |
| 36-50 | 4.260 | 3.170 (74) |
| 51-66 | 2.610 | 2.090 (80) |
| 67-79 | 720 | 590 (83) |
| 80+ | 70 | 50 (76) |
| **Voting proportions political movements (%)** | | |
| **Progressive liberal** | | |
| 0-0.1 | 1.458.190 | 1.113.830 (76) |
| 18-35 | 386.960 | 237.550 (61) |
| 36-50 | 328.030 | 240.330 (73) |
| 51-66 | 413.890 | 347.260 (84) |
| 67-79 | 239.630 | 213.130 (89) |
| 80+ | 89.690 | 75.550 (84) |
| 0.1-0.2 | 8.902.660 | 7.139.080 (80) |
| 18-35 | 2.301.410 | 1.493.560 (65) |
| 36-50 | 2.031.010 | 1.570.240 (77) |
| 51-66 | 2.530.710 | 2.211.210 (87) |
| 67-79 | 1.481.490 | 1.365.810 (92) |
| 80+ | 558.040 | 498.270 (89) |
| 0.2-0.3 | 2.635.610 | 2.126.450 (81) |
| 18-35 | 795.040 | 551.320 (69) |
| 36-50 | 642.250 | 512.170 (80) |
| 51-66 | 666.320 | 581.160 (87) |
| 67-79 | 373.450 | 341.140 (91) |
| 80+ | 158.560 | 140.660 (89) |
| 0.3-0.4 | 1.042.600 | 828.770 (79) |
| 18-35 | 453.500 | 340.020 (75) |
| 36-50 | 235.450 | 184.540 (78) |
| 51-66 | 211.780 | 179.520 (85) |
| 67-79 | 105.840 | 93.840 (89) |

| | | |
|---|---|---|
| 80+ | 36.030 | 30.850 (86) |
| 0.4-0.5 | 135.420 | 114.110 (84) |
| 18-35 | 74.860 | 61.170 (82) |
| 36-50 | 23.070 | 19.240 (83) |
| 51-66 | 22.690 | 20.160 (89) |
| 67-79 | 11.440 | 10.570 (92) |
| 80+ | 3.360 | 2.980 (89) |
| Unknown | 1.160 | 560 (48) |
| 18-35 | 560 | 210 (38) |
| 36-50 | 310 | 150 (48) |
| 51-66 | 220 | 150 (66) |
| 67-79 | 70 | 50 (78) |
| 80+ | 10 | . |
| **Right-wing liberal** | | |
| 0-0.1 | 373.280 | 245.010 (66) |
| 18-35 | 157.910 | 94.170 (60) |
| 36-50 | 91.290 | 58.280 (64) |
| 51-66 | 80.420 | 58.910 (73) |
| 67-79 | 33.880 | 26.410 (78) |
| 80+ | 9.790 | 7.240 (74) |
| 0.1-0.2 | 5.449.320 | 4.123.990 (76) |
| 18-35 | 1.797.910 | 1.149.040 (64) |
| 36-50 | 1.231.000 | 899.710 (73) |
| 51-66 | 1.353.570 | 1.132.140 (84) |
| 67-79 | 771.610 | 688.840 (89) |
| 80+ | 295.230 | 254.260 (86) |
| 0.2-0.3 | 7.101.440 | 5.876.040 (83) |
| 18-35 | 1.764.450 | 1.222.390 (69) |
| 36-50 | 1.638.170 | 1.316.970 (80) |
| 51-66 | 2.042.290 | 1.812.370 (89) |
| 67-79 | 1.197.490 | 1.112.150 (93) |
| 80+ | 459.050 | 412.160 (90) |
| 0.3-0.4 | 1.244.820 | 1.072.320 (86) |
| 18-35 | 290.730 | 217.470 (75) |
| 36-50 | 298.130 | 250.530 (84) |
| 51-66 | 367.320 | 334.310 (91) |
| 67-79 | 207.720 | 196.010 (94) |
| 80+ | 80.920 | 74.000 (91) |
| 0.4-0.5 | 5.620 | 4.880 (87) |
| 18-35 | 780 | 540 (70) |
| 36-50 | 1.220 | 1.030 (85) |
| 51-66 | 1.780 | 1.580 (89) |
| 67-79 | 1.160 | 1.080 (93) |

| | | |
|---|---|---|
| 80+ | 690 | 650 (93) |
| Unknown | 1.160 | 560 (48) |
| 18-35 | 560 | 210 (38) |
| 36-50 | 310 | 150 (48) |
| 51-66 | 220 | 150 (66) |
| 67-79 | 70 | 50 (78) |
| 80+ | 10 | . |
| **Right-wing Christian** | | |
| 0-0.1 | 13.501.210 | 10.833.030 (80) |
| 18-35 | 3.821.890 | 2.574.950 (67) |
| 36-50 | 3.107.460 | 2.419.810 (78) |
| 51-66 | 3.662.560 | 3.190.390 (87) |
| 67-79 | 2.105.720 | 1.933.000 (92) |
| 80+ | 803.590 | 714.880 (89) |
| 0.1-0.2 | 359.010 | 278.500 (78) |
| 18-35 | 95.340 | 59.940 (63) |
| 36-50 | 81.040 | 60.680 (75) |
| 51-66 | 98.680 | 83.980 (85) |
| 67-79 | 59.720 | 53.500 (90) |
| 80+ | 24.230 | 20.400 (84) |
| 0.2-0.3 | 169.960 | 120.090 (71) |
| 18-35 | 49.670 | 27.450 (55) |
| 36-50 | 38.160 | 26.120 (68) |
| 51-66 | 45.170 | 36.170 (80) |
| 67-79 | 26.330 | 22.290 (85) |
| 80+ | 10.630 | 8.050 (76) |
| 0.3-0.4 | 48.250 | 31.370 (65) |
| 18-35 | 15.310 | 7.960 (52) |
| 36-50 | 11.710 | 7.380 (63) |
| 51-66 | 12.510 | 9.440 (75) |
| 67-79 | 6.400 | 5.020 (78) |
| 80+ | 2.330 | 1.570 (67) |
| 0.4-0.5 | 42.880 | 23.380 (55) |
| 18-35 | 14.350 | 5.820 (41) |
| 36-50 | 9.720 | 5.120 (53) |
| 51-66 | 10.860 | 7.110 (66) |
| 67-79 | 5.730 | 4.020 (70) |
| 80+ | 2.220 | 1.320 (59) |
| 0.5-0.6 | 21.750 | 8.720 (40) |
| 18-35 | 8.030 | 2.040 (25) |
| 36-50 | 5.190 | 1.970 (38) |
| 51-66 | 5.160 | 2.770 (54) |
| 67-79 | 2.530 | 1.540 (61) |

| | | |
|---|---|---|
| 80+ | 830 | 400 (48) |
| 0.7+ | 400 | 160 (39) |
| 18-35 | 180 | 50 (26) |
| 36-50 | 100 | 40 (43) |
| 51-66 | 100 | 50 (50) |
| 67-79 | 20 | 10 (58) |
| 80+ | 10 | . |
| Unknown | 32.180 | 27.550 (86) |
| 18-35 | 7.570 | 5.620 (74) |
| 36-50 | 6.720 | 5.550 (82) |
| 51-66 | 10.580 | 9.540 (90) |
| 67-79 | 5.460 | 5.150 (94) |
| 80+ | 1.850 | 1.690 (91) |
| **Christian middle** | | |
| 0-0.1 | 5.671.140 | 4.325.370 (76) |
| 18-35 | 1.899.850 | 1.228.340 (65) |
| 36-50 | 1.349.300 | 1.002.580 (74) |
| 51-66 | 1.401.260 | 1.184.530 (85) |
| 67-79 | 742.270 | 666.560 (90) |
| 80+ | 278.450 | 243.370 (87) |
| 0.1-0.2 | 6.818.410 | 5.633.990 (83) |
| 18-35 | 1.674.670 | 1.157.410 (69) |
| 36-50 | 1.532.780 | 1.228.230 (80) |
| 51-66 | 1.965.300 | 1.738.410 (88) |
| 67-79 | 1.186.740 | 1.099.500 (93) |
| 80+ | 458.920 | 410.440 (89) |
| 0.2-0.3 | 1.371.420 | 1.103.080 (80) |
| 18-35 | 355.040 | 238.790 (67) |
| 36-50 | 307.190 | 239.030 (78) |
| 51-66 | 389.670 | 337.710 (87) |
| 67-79 | 230.780 | 210.420 (91) |
| 80+ | 88.740 | 77.150 (87) |
| 0.3-0.4 | 276.010 | 229.180 (83) |
| 18-35 | 71.300 | 51.340 (72) |
| 36-50 | 61.410 | 49.410 (80) |
| 51-66 | 79.080 | 69.780 (88) |
| 67-79 | 46.480 | 42.900 (92) |
| 80+ | 17.740 | 15.760 (89) |
| 0.4-0.5 | 37.490 | 30.600 (82) |
| 18-35 | 10.910 | 7.730 (71) |
| 36-50 | 9.110 | 7.280 (80) |
| 51-66 | 10.080 | 8.890 (88) |
| 67-79 | 5.570 | 5.110 (92) |

| | | |
|---|---|---|
| 80+ | 1.830 | 1.600 (88) |
| Unknown | 1.160 | 560 (48) |
| 18-35 | 560 | 210 (38) |
| 36-50 | 310 | 150 (48) |
| 51-66 | 220 | 150 (66) |
| 67-79 | 70 | 50 (78) |
| 80+ | 10 | . |
| **Right-wing conservative** | | |
| 0-0.1 | 1.122.670 | 875.120 (78) |
| 18-35 | 479.340 | 350.980 (73) |
| 36-50 | 254.890 | 194.940 (76) |
| 51-66 | 232.860 | 193.770 (83) |
| 67-79 | 115.110 | 100.800 (88) |
| 80+ | 40.480 | 34.630 (86) |
| 0.1-0.2 | 5.939.710 | 4.798.900 (81) |
| 18-35 | 1.691.030 | 1.157.600 (68) |
| 36-50 | 1.388.260 | 1.096.670 (79) |
| 51-66 | 1.576.110 | 1.378.350 (87) |
| 67-79 | 916.020 | 840.240 (92) |
| 80+ | 368.290 | 326.060 (89) |
| 0.2-0.3 | 6.238.150 | 4.973.740 (80) |
| 18-35 | 1.621.070 | 1.043.330 (64) |
| 36-50 | 1.423.280 | 1.093.280 (77) |
| 51-66 | 1.777.640 | 1.547.310 (87) |
| 67-79 | 1.032.020 | 948.690 (92) |
| 80+ | 384.140 | 341.130 (89) |
| 0.3-0.4 | 815.300 | 631.040 (77) |
| 18-35 | 205.270 | 122.990 (60) |
| 36-50 | 179.320 | 131.760 (73) |
| 51-66 | 241.060 | 205.290 (85) |
| 67-79 | 139.510 | 126.700 (91) |
| 80+ | 50.140 | 44.300 (88) |
| 0.4-0.5 | 32.580 | 23.490 (72) |
| 18-35 | 8.310 | 4.430 (53) |
| 36-50 | 7.940 | 5.470 (69) |
| 51-66 | 10.210 | 8.300 (81) |
| 67-79 | 4.730 | 4.140 (88) |
| 80+ | 1.390 | 1.150 (83) |
| 0.5-0.6 | 26.080 | 19.930 (76) |
| 18-35 | 6.750 | 4.280 (63) |
| 36-50 | 6.100 | 4.410 (72) |
| 51-66 | 7.500 | 6.290 (84) |
| 67-79 | 4.470 | 3.910 (87) |

| | | |
|---|---|---|
| 80+ | 1.250 | 1.040 (83) |
| Unknown | 1.160 | 560 (48) |
| 18-35 | 560 | 210 (38) |
| 36-50 | 310 | 150 (48) |
| 51-66 | 220 | 150 (66) |
| 67-79 | 70 | 50 (78) |
| 80+ | 10 | . |
| **Progressive left-wing** | | |
| 0-0.1 | 467.900 | 339.290 (73) |
| 18-35 | 137.840 | 80.980 (59) |
| 36-50 | 107.150 | 74.860 (70) |
| 51-66 | 125.820 | 102.030 (81) |
| 67-79 | 69.990 | 59.950 (86) |
| 80+ | 27.100 | 21.480 (79) |
| 0.1-0.2 | 5.206.530 | 4.340.030 (83) |
| 18-35 | 1.272.160 | 895.490 (70) |
| 36-50 | 1.218.320 | 988.790 (81) |
| 51-66 | 1.516.920 | 1.350.910 (89) |
| 67-79 | 870.730 | 810.120 (93) |
| 80+ | 328.410 | 294.730 (90) |
| 0.2-0.3 | 6.603.020 | 5.195.710 (79) |
| 18-35 | 1.839.480 | 1.175.010 (64) |
| 36-50 | 1.509.220 | 1.147.940 (76) |
| 51-66 | 1.783.490 | 1.539.630 (86) |
| 67-79 | 1.055.650 | 965.120 (91) |
| 80+ | 415.190 | 368.020 (89) |
| 0.3-0.4 | 1.660.520 | 1.278.500 (77) |
| 18-35 | 674.370 | 476.470 (71) |
| 36-50 | 367.600 | 274.760 (75) |
| 51-66 | 359.590 | 299.830 (83) |
| 67-79 | 189.950 | 168.110 (89) |
| 80+ | 69.000 | 59.320 (86) |
| 0.4-0.5 | 236.510 | 168.710 (71) |
| 18-35 | 87.930 | 55.670 (63) |
| 36-50 | 57.520 | 40.190 (70) |
| 51-66 | 59.570 | 46.910 (79) |
| 67-79 | 25.530 | 21.180 (83) |
| 80+ | 5.980 | 4.770 (80) |
| Unknown | 1.160 | 560 (48) |
| 18-35 | 560 | 210 (38) |
| 36-50 | 310 | 150 (48) |
| 51-66 | 220 | 150 (66) |
| 67-79 | 70 | 50 (78) |

| | | |
|---|---|---|
| 80+ | 10 | . |

### S4 Vaccine uptake in the Netherlands per neighbourhood

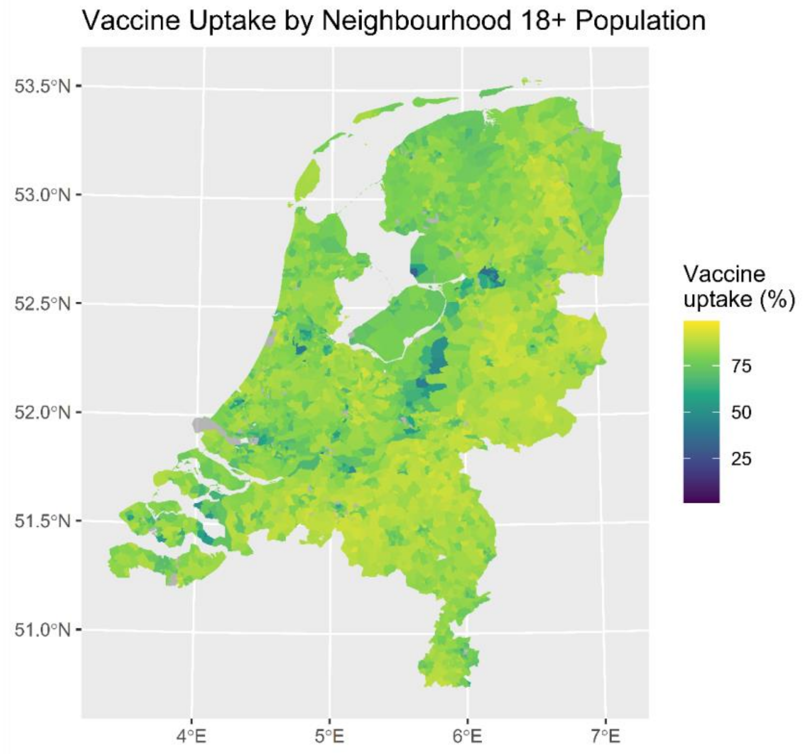

**Figure S4.1 Vaccine uptake (%) in the Netherlands per neighbourhood in the population of 18 years and older**

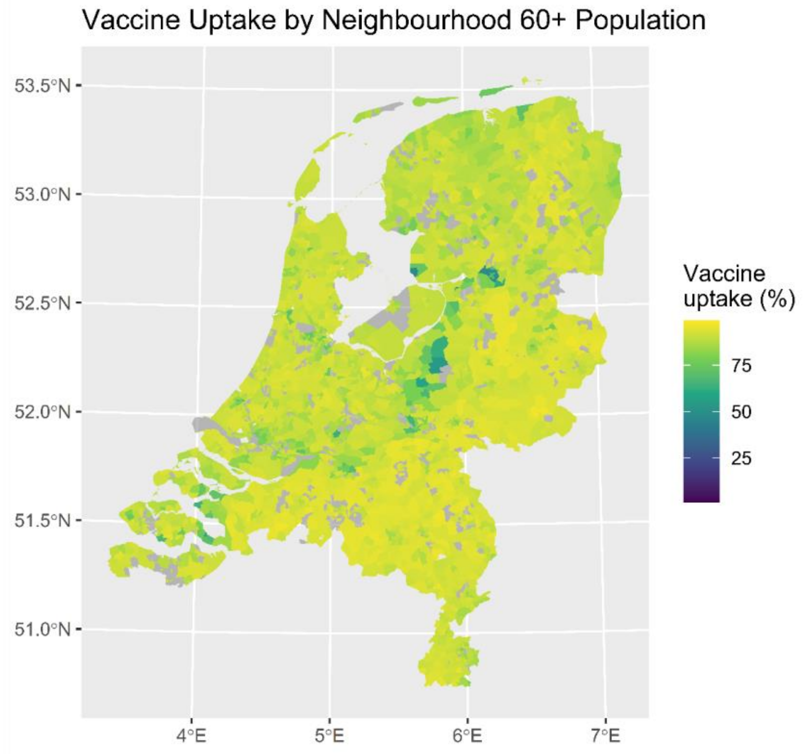

**Figure S4.2 Vaccine uptake (%) in the Netherlands per neighbourhood in the population**

**of 60 years and older**
Note: the grey areas indicate neighbourhoods with frequencies <10 and were therefore excluded
